# Clinical Knowledge and Reasoning Abilities of AI Large Language Models in Anesthesiology: A Comparative Study on the ABA Exam

**DOI:** 10.1101/2023.05.10.23289805

**Authors:** Mirana C. Angel, Joseph B. Rinehart, Maxime P. Canneson, Pierre Baldi

## Abstract

Over the past decade, Artificial Intelligence (AI) has expanded significantly with increased adoption across various industries, including medicine. Recently, AI’s large language models such as GPT-3, Bard, and GPT-4 have demonstrated remarkable language capabilities. While previous studies have explored their potential in general medical knowledge tasks, here we assess their clinical knowledge and reasoning abilities in a specialized medical context. We study and compare their performances on both the written and oral portions of the comprehensive and challenging American Board of Anesthesiology (ABA) exam, which evaluates candidates’ knowledge and competence in anesthesia practice. In addition, we invited two board examiners to evaluate AI’s answers without disclosing to them the origin of those responses. Our results reveal that only GPT-4 successfully passed the written exam, achieving an accuracy of 78% on the basic section and 80% on the advanced section. In comparison, the less recent or smaller GPT-3 and Bard models scored 58% and 47% on the basic exam, and 50% and 46% on the advanced exam, respectively. Consequently, only GPT-4 was evaluated in the oral exam, with examiners concluding that it had a high likelihood of passing the actual ABA exam. Additionally, we observe that these models exhibit varying degrees of proficiency across distinct topics, which could serve as an indicator of the relative quality of information contained in the corresponding training datasets. This may also act as a predictor for determining which anesthesiology subspecialty is most likely to witness the earliest integration with AI.

## Introduction

In recent years, Artificial Intelligence (AI) primarily in the form of machine learning, in particular deep learning, has experienced a significant expansion driven primarily by progress in computational power and big data availability.^1^ In the medical field, AI’s potential to increase accuracy and expedite diagnoses has led to its application in numerous areas, including radiology, pathology, and genomics. For example, AI has been employed to examine medical images and videos for early indicators of diseases such as cancer, predict disease likelihood, and tailor treatment plans based on a patient’s genetic profile.^2,3,4^ In addition, AI-based large language models (LLMs), trained on very large corpora of text, are now able to fluidly generate high-quality text (and software) on almost any subject, opening new opportunities for transforming healthcare. But how knowledgeable and capable of reasoning are LLMs in medicine? One previous study has explored their potential in general medical knowledge tasks.^5^ Here we assess their clinical knowledge and reasoning abilities in a specialized medical context.

Most LLMs utilize neural network architectures known as transformers, which incorporate attention mechanisms for processing input sequences. This approach has demonstrated significant efficacy in tasks such as language translation and text generation.^6,7^ Here we consider some of the currently most notable LLMs, including the Generative Pre-trained Transformer-3 (GPT-3), Bard, and the Generative Pre-trained Transformer-4 (GPT-4). ^8,9,10^

These models utilize the transformer’s decoder-only architecture^9,11^, and their primary differences lie in their size and the nature of the data they were trained on. The GPT-3 model possesses 175 billion parameters and has already been shown to excel in several tasks. The GPT-4 model overcomes many of the limitations of GPT-3 by enlarging the model size to one trillion parameters. Both versions of GPT were pre-trained on a vast text corpus, followed by fine-tuning to perform specific tasks.^8,9^ In addition, the models were enhanced using the Reinforcement Learning from Human Feedback (RLHF) method, which utilizes human-generated feedback to enable model alignment to human preferences, guiding the model toward generating more accurate and appropriate responses.^10^ On the other hand, Google’s Bard employs the Language Model for Dialogue Applications (LaMDA) with 137 billion parameters and is primarily pre-trained on public dialog data and web text, which leads to improved conversational understanding and accurate dialog-style responses.^11^

These systems, which are more or less capable of passing the Turing test and far exceed humans in their speed, fluidity, and capability to speak many languages, raise the anthropomorphic, ill-defined, question of how much LLMs can actually “understand”. Many studies suggest that LLMs are prone to hallucinations and errors, sometimes struggling with reasoning, causality, and common-sense understanding.^12,13^ However, it is important to acknowledge that complex systems operating in large domains are bound to make errors occasionally, similar to visual illusions in the human visual cortex. Moreover, LLMs have been rapidly improving, with fewer and fewer errors being observed, and it has been shown that careful prompting can aid LLMs in focusing, self-correcting, and reasoning, improving their capabilities even further. ^14^

To assess the capability of LLMs to understand medical data, researchers recently evaluated GPT-3’s capacity to answer questions from the United States Medical Licensing Examination (USMLE). The study’s results demonstrated that the model performed exceptionally well on the exam, exhibiting a high degree of accuracy and fluency in medical reasoning.^15^ Thus these initial results suggest that large language models have the potential for transforming both medical education and practice.

While the USMLE is a widely recognized exam for general medical knowledge, it may not fully encompass the complex scenarios medical specialists face in their practice. Hence, in this work, we aim to further assess the clinical knowledge and reasoning abilities of large language models, namely GPT-3, BARD, and GPT-4, by examining their performance on the American Board of Anesthesiology (ABA) exam. This exam comprises three parts: the Basic Exam, the Advanced Exam, and the Applied Exam. The Basic Exam evaluates fundamental knowledge in anesthesiology and is typically taken during the second post-graduation year (PGY-2) of residency training. The Advanced Exam is more comprehensive, covering topics such as patient care, pharmacology, and medical knowledge, and is taken in the first year following graduation from a residency program. The Applied Exam, taken only after passing the first two exams, finally assesses a candidate’s clinical competence in anesthesia practice, comprising both a Structured Oral Examination (SOE) and a hands-on Objective Structured Clinical Examination (OSCE). The tri-part ABA exam is considered one of the most challenging exams in the medical field, necessitating extensive knowledge and training in anesthesiology. Passing the exams is a significant accomplishment and is required for certification by the American Board of Anesthesiology.^16,17^ By analyzing the models’ performance on this exam, we can better comprehend their potential utility in highly specialized and versatile medical scenarios, offering insights into the strengths and limitations of large language models in the medical domain.

## Material and Methods

The ABA examination is a tripartite evaluation that encompasses the Basic Exam, Advanced Exam, and Applied Exam. For the basic exam, we used the full set of sample questions provided on the ABA website, which contains 60 multiple-choice questions with answer keys provided.^18^

The Advanced Exam was formulated using the book, “Anesthesia Review: 1000 Questions and Answers to Blast the BASICS and Ace the ADVANCED.” This book’s advanced topics section comprises 14 detailed chapters in the following domains: Advanced Monitors, Pain, Pediatric Anesthesia, Obstetric Anesthesia, Cardiac and Vascular Anesthesia, Thoracic Anesthesia, ENT Anesthesia, Anesthesia for Special Indications, Orthopedic Anesthesia, Trauma, Anesthesia for Ambulatory Surgery, Geriatrics, Critical Care, and Ethics.^19^ We chose five questions at random from each chapter, resulting in a final 70-item multiple-choice questionnaire. The multiple-choice questions and possible answers from both the basic and advanced sample exams were entered into GPT-3 and GPT-4 using the ChatGPT plus user interface^20^ and entered into Bard using the user interface provided by Google.^21^ The questions were entered individually and the answer from the AI was recorded. Some of the multiple-choice questions contained images that could not serve as input for these language models. In these cases, we described the images using words. The selected multiple-choice responses from the models were finally compared with the answer keys for scoring.

For the Applied exam, it was not feasible to perform the OSCE portion of the advanced exam which involves things like hands-on ultrasound stations and interpretation of live monitors, so we were only able to explore the SOE component (we assume at the present time no AI is prepared to pass the interactive OSCE). Further, we only tested GPT-4 in the SOE because GPT-3 and Bard did not pass the basic and advanced multiple-choice exams.

The SOE exam is described by the American Society of Anesthesiologists on their website as being intended “to assess your judgment, adaptability, and organization.” As such, the examiners are trained to probe examinees for both depth and breadth of knowledge, assess adaptability in light of new information, and do so in a very time-intensive condition. This exam format and the interactive cadence under time pressure are nearly impossible to replicate faithfully with a typed terminal interaction, and each one of the examiners who reviewed the blinded results individually commented that their ability to “assess” a candidate was extremely limited without the ability to interact directly themselves under the true testing conditions. Further, as an AI model, GPT-4 has no true “Judgment” or “Adaptability” (or even true application of information) in the form we think of to be tested. Nevertheless, the content of the responses can certainly be judged for organization and the *appearance* or presentation of judgment and adaptability, within the noted limitations. The other challenge in interpreting the results is that for the SOE each examinee is scored by multiple examiners, and each individual score is normalized for the examiner’s specific scoring style as well as the scores for that specific SOE question at that day and time and more, all of which make it nearly impossible for any examiner to definitively and solely judge “passing” versus “failing”. Thus, the most objective comparison we could hope to achieve was to solicit examiner *opinions* on whether the observed performance was high or low in their opinion, and whether they believed in their experience this was likely passing, failing, or indeterminate performance.

For the SOE stem we used the sample exam found on the ABA website. We chose the first long-stem format which includes sub-topics in intraoperative (4 questions) and post-operative care (6 questions), followed by three unrelated patient short-stem topics.^22^ ChatGPT-4, even under the paid subscription model, currently has a limit of ‘questions’ that can be asked in a day. The interactive nature of the SOE caused that limit to be hit quickly in just a fraction of the total exam, so we had to be strategic in our interaction with the AI model. We chose, therefore, to ‘administer’ the exam in two distinct approaches. In the first approach, we provided the AI with the complete patient stem, and then presented the entirety of a question block at once and allowed the AI to respond to the entire block at once. While dissimilar to the actual exam format, this allowed us to reduce the number of “questions” being counted by the AI’s daily limit and assess the complete responses of the AI to the exam topics for the component when presented in their entirety.

In the second format, we used an interactive approach to the questions more consistent with the actual exam format, spreading the interactions out over several days when the ceiling was hit each day. The exam was managed by an ABA examiner and included additional attempts for probing depth, breadth, adaptability, and detail as might be encountered during the examination. In particular, we focused on the application of knowledge and decision-making (judgment) in responses in this portion (which we assumed might be more challenging) as opposed to medical knowledge and organization (which we assumed would be easier for the AI).

Since the question stem we used was found on the ABA website it was possible that written “ideal’ responses might exist somewhere on the web that had been incorporated into the AI database, so to pursue the interactive format in greater depth and novelty, a different “grab-bag” topic stem was presented from a very old exam (one of the authors’ favorites for really challenging critical thinking and application) and pursued with a bit more novelty and open exploration, asking the AI to make medical judgment calls in a very brief but potentially complex and unstated case history.

For the purposes of this exam, any qualifications the AI made about being an AI were ignored (and deleted when shared with the blinded reviewers), and when possible firm answers were solicited. The AI model was given instructions at the beginning of the exam to “pretend it was being examined in anesthesiology”, that it was the highest-level expert available, and to keep its responses as terse as possible as though it was being examined in a timed test while retaining the highest priority information in the responses.

For the SOE exam scoring, two authors – one a current ABA board examiner (JR) and one the chair of his department and an experienced clinician (MC) – evaluated the AI responses in comparison to their years of experience with actual examinees and trainees and rated the responses on their approximated probability that a human respondent would pass the exam if providing the given performance. As a further validation, the second exam format was shared with two other current or former ABA SOE examiners who were blinded to the origin of the responses and asked to review a portion of “a transcript from a mock oral exam to be used for resident training purposes” and provide their thoughts on whether they thought this mock examinee’s performance was a good example or not, and whether they would ultimately pass if this was their true exam.

## Results

In the assessment with basic exam questions, GPT-3 and Bard achieved scores of 58.33% and 46.67% respectively, which suggests that they would likely be unable to pass a comprehensive 200-question examination. In contrast, the more advanced language model, GPT-4, obtained a score of 78.33%, demonstrating a greater probability of successfully passing the actual examination (Table 1).

**Table 1:** Test Results of Different Models on the Sample Basic Exam

| Model | Raw Score | Percent Score |
| --- | --- | --- |
| GPT-3 | 35/60 | 58.33% |
| Bard | 28/60 | 46.67% |
| GPT-4 | 47/60 | 78.33% |

In the advanced assessment sample, GPT-3 and Bard obtained scores of 50.00% and 45.71% respectively, suggesting that their chances of passing a comprehensive 200-question examination are low. However, the more sophisticated language model, GPT-4, achieved an 80.00% score, surpassing its performance in the basic exam and demonstrating a higher probability of successfully completing the actual advanced examination (Table 2). Moreover, the performance of the models varied across individual subtopics as illustrated in Table 2 and Figure 1.

**Table 2:** Test Result of the Sample Advanced Exam of Different Models by Topic

| Topics | GPT-3 | Bard | GPT-4 |
| --- | --- | --- | --- |
| Advanced Monitors | 3/5 | 3/5 | 4/5 |
| Pain | 5/5 | 5/5 | 5/5 |
| Pediatric Anesthesia | 3/5 | 1/5 | 3/5 |
| Obstetric Anesthesia | 1/5 | 1/5 | 3/5 |
| Cardiac and Vascular Anesthesia | 2/5 | 4/5 | 4/5 |
| Thoracic Anesthesia | 5/5 | 5/5 | 5/5 |
| ENT Anesthesia | 2/5 | 2/5 | 4/5 |
| Anesthesia for Special Indications | 3/5 | 4/5 | 4/5 |
| Orthopedic Anesthesia | 0/5 | 0/5 | 2/5 |
| Trauma | 1/5 | 0/5 | 3/5 |
| Anesthesia for Ambulatory Surgery | 2/5 | 0/5 | 4/5 |
| Geriatrics | 3/5 | 2/5 | 5/5 |
| Critical Care | 2/5 | 2/5 | 5/5 |
| Ethics and Professionalism | 3/5 | 3/5 | 5/5 |
| Total Raw Score | 35/70 | 32/70 | 56/70 |
| Total Percent Score | 50.00% | 45.71% | 80.00% |

**Figure 1:**
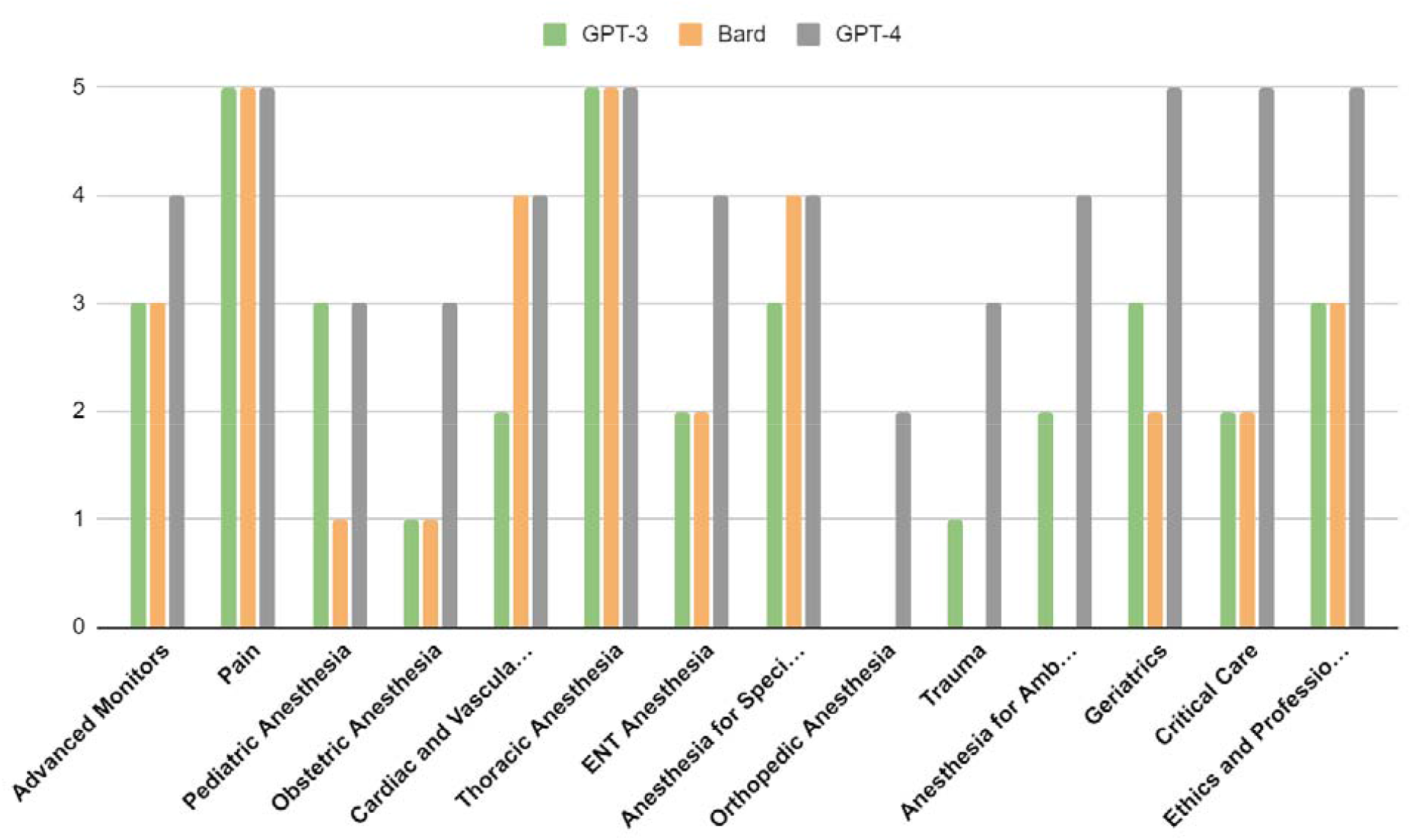
Number of Correct Responses per Five Questions in each Topic in the Sample Advanced Produced by Different Models

The SOE interactions are provided as Appendix 1 (the static exam with complete questions asked en bloc) and Appendix 2 (the interactive format). On the SOE portion of the applied exam, while not consistent with an actual exam for the “complete question and answers” format initially tried, both evaluators agreed the contents of the responses had some notable gaps but did not raise any critical “failing” concerns. The organization of the responses was, unsurprisingly, excellent. The medical knowledge content was also high as might be expected. The incorporation of information about the current patient was also quite good, with the AI consistently incorporating specific patient details into the responses it provided and the “decision-making” descriptions. During the response to hemorrhage, for example, the AI specifically stated it would “administer the two units that are available while asking for additional units”. In general, the incorporation of information from the patient stem was above what might be expected during a real exam, and the blood gas analysis was excellent. The AI response even appears to demonstrate judgment via risk assessment when, in response to differences in extubation criteria for this thoracotomy patient compared to a cholecystectomy patient, it states: “In an ASA-1 cholecystectomy patient, these specific concerns might not be as significant, as they typically have no significant medical history and are undergoing a less invasive surgery.”

In the ‘interactive’ format, a few more deficiencies were found with the AI responses, and repetition of some phrases and elements was seen, though this latter may have been modifiable with additional instructions to the AI to limit repetition. Both unblinded evaluators considered the responses sub-ideal, but potentially passing if it had come from a true exam. Asking for clarifications or asking for additional depth, changing the scenario, and even asking the AI to prioritize importance when it gave detailed lists resulted in generally reasonable responses.

The deficiencies most likely encountered were improper prioritization and inappropriate choices. For example, the AI did not focus on the current neurological and vascular status of the carotid endarterectomy patient to understand the urgency of a new procedure, focusing instead on anesthetic and medication history (Appendix 2 lines 377-407). Another example is the AI initially maintaining that a left-sided double-lumen tube would be preferred even for a left pneumonectomy procedure (Appendix 2 lines 97-101). There was additional confusion around optimizing oxygenation in one-lung ventilation in the responses (Appendix 2 lines 174-283). When these possibly concerning responses were explored, however, as an ABA examiner would do during the actual exam, the AI in each case ultimately ended up appropriately revising its initial flawed decisions. After questioning a left sided-tube in the presence of a hilar resection, the AI “realized” a left-sided tube was problematic and then stated it would revise its previous response (Appendix 2 lines 101-117). It did the same for the confused portions of oxygenation in one-lung ventilation (Appendix 2 lines 232-281), and the neurological history of the CEA patient (Appendix 2 lines 406-420). During the testing, we began to wonder if the AI was simply changing priorities in response to any subtle suggestion to do so, so we deliberately followed the same questioning format for what we considered a moderate but lower priority history for the CEA patient: renal function. Interestingly, in this circumstance the AI correctly described the importance of this system but chose *not* to revise its previous response when asked (appropriately in our opinion), suggesting there is some decision function taking place and it isn’t just responding to the user input as cues (Appendix 2 lines 438-466).

The unblinded evaluators both noted that the evaluation of the SOE exam is complex. Real candidates make simple and even large mistakes frequently during sessions due to the stress, importance, and speed of the exam format and the examiners usually choose to probe such mistakes a bit to give the examinee time to reconsider their responses. As such, while a mistake followed by a correction on exploration is obviously less preferred than an initially correct response, such mistakes are not usually ‘failing criteria’ when corrected, and even if uncorrected may not be sufficient alone if performance otherwise is good. In the present case, the evaluators felt that the mistakes being made were of moderate concern, but with appropriate corrections when probed. A human candidate responding in this manner would not receive a definite failure, but neither would they receive a definite pass. Beyond that, it is impossible to be certain, but both unblinded reviewers guessed that the performance, as observed, was probably more likely to pass than not. Certainly, the exceptional organization and clarity of the responses (which *were* superior to most true examinees) would have benefitted the exam.

The blinded reviewers also both immediately qualified that interpretation of performance via a written transcript, without the ability to interact and the experience of the time pressure during the exam, was limited. They all also commented that, despite the attempts to instruct the AI to limit responses, the “examinee” responses were too wordy and often repetitive. Both blinded reviewers, in light of the limitations, expressed that while the performance was ‘moderate’ there was ‘some’ to ‘reasonable’ probability that the examinee would ultimately pass the exam.

## Discussion

The passing threshold for the ABA written exam varies annually, however, it is reasonable to posit that a score exceeding 75% typically denotes a passing grade.^23^ Consequently, only GPT-4 managed to pass both the basic and advanced exams. This finding further corroborates the hypothesis that enhancing model size can improve task performance.^8,9^ As future language models become increasingly large and intricate, it is plausible to anticipate accuracy levels nearing 100% when evaluating them with analogous exam problems.

While GPT-3 and Bard were unable to pass the multiple-choice exams, examining their performance across various topics remains intriguing. For the advanced exam sample, we randomly selected practice questions from 14 distinct topics, with the models displaying differential performance in each. Remarkably, all models obtained perfect scores in thoracic and pain management categories, while they achieved the lowest scores in obstetrics, orthopedics, trauma, and ambulatory surgeries (Figure 1). Given that the models’ training data predominantly originates from the internet, this suggests a correlation between the availability of accurate information on specific topics and the models’ performance. Furthermore, these findings may serve as an indicator for prioritizing AI integration within certain anesthesia subspecialties.

Furthermore, we observed that the language models exhibit a substantially higher accuracy when presented with multiple-choice questions involving purely textual content. However, their accuracy declines significantly when faced with questions containing numerical calculations or involving numbers. This can be attributed to the fact that these language models are predominantly trained on text-based data and lack specific training in numerical calculations and the language constructs around conveying these calculations.^7,8,9,11^ As a result, the models struggle to fully comprehend questions involving numerical calculations, leading to imprecise responses. Additionally, numerical calculations demand greater precision and exactness, which may not always be within the model’s capabilities, resulting in further inaccuracies. To enhance the accuracy of large language models in solving problems with numerical calculations, it is essential to incorporate a specialized training dataset comprising numerical data during the model’s training phase.

The applied SOE exam results are perhaps more impressive than the multiple-choice exam performance. While only GPT-4 was tested at this level, the evaluators felt that the responses and performance were ‘moderate’ when compared to an anesthesiology residency-trained physician who had already passed the written exams. When unblinded and understood to come from an AI model that didn’t go to medical school or residency or have any experience actually sitting in an operating room and managing an anesthetic, however, the responses to the questions were incredibly concise, informative, and inclusive of the patient background history provided. The adaptation of responses to new information was convincing, as was the revision of previous statements considering subsequent questions.

As noted above, the only real initial “flaw” we detected during examining was the prioritization of information when asked to narrow down a long list (which in all fairness the AI did revise later when asked to reconsider), and apparent gaps in the information incorporated in an initial response. Based on the known limitations of these AI models at this stage, it seems plausible that limits are more likely to be found with the more synthetic (and less “regurgitative”) exams, but even this is not certain as overall the AI’s “synthetic” functions in replicating the application and presentation of judgment were certainly superior to at least some human examinees we have encountered. Moreover, as these models continue to expand in training data, size, and scope, there is no reason to expect that performance will do anything but improve.

Interpreting the results from the standpoint of the AI itself, the *context* of the responses was always accurate - the AI always appeared to attempt to answer the intent of the question, demonstrating the impressive natural language processing of the system. Anecdotally, ‘mistakes’ seemed to come from ‘blind spots’ in the information incorporated into the responses. When the blind spot was probed and the AI focused on it, the subsequent responses improved. If we were going to engage in a series of such exams in written format with human and AI respondents, our best guess to differentiate the two would be to ask broad questions that require large amounts of unstated assumptions to be incorporated in order to provide a correct response. We would then attempt to identify the humans via the inclusion, and the machines via the inclusion gaps, of all relevant, but especially ‘obvious’ circumstantial unstated data. For example, that a left pneumonectomy requires hilar resection, and a left-sided tube is not feasible, is something that *should* be obvious to trained anesthesiologists, but the AI did not seem to incorporate it until it was asked to focus on exactly that point.

As it stands, the performance of the AI in this process was impressive enough that it leads one to question whether we shouldn’t already, as highly trained clinicians, be using these AI systems to help us avoid some of the more common cognitive errors that may occur during critical events. We have a code leader at all hospital codes, and typically a dedicated scribe to record events. Perhaps an additional resource to consult the AI for missed items on the differential and missed priority interventions to prompt the humans in the room is a well-invested effort.

The ability of GPT-4 to pass the ABA examination carries both positive and negative implications for medical education. On one hand, the model’s success highlights the potential of large language models to act as influential tools in medical education, enhancing the quality of instruction and preparing students for examinations. This progress could pave the way for the creation of more efficient and effective pedagogical approaches, offering students tailored learning experiences that accommodate their unique learning styles.

On the other hand, there are potential negative consequences that educators must consider. The overreliance on large language models could contribute to a decline in critical thinking and problem-solving abilities, as students might depend excessively on the model’s outputs rather than cultivating their own analytical skills. To address this, educators could assign tasks involving greater reasoning and numerical analysis, and place increased emphasis on in-person discussions over written assignments. In summary, the incorporation of large language models into medical education holds the potential to transform the way students learn; however, educators must remain cognizant of their limitations and strive to maintain a balance with traditional teaching methods.

## Conclusion

This study evaluated the clinical knowledge and reasoning capabilities of large language models, specifically GPT-4, in anesthesiology by assessing their performance on the American Board of Anesthesiology exam. Our findings highlight the relationship between model size and task accuracy and suggest potential areas for AI integration within anesthesiology subspecialties based on varied performance across topics. Future research should address the limitations and ethical implications of deploying AI in clinical settings, as well as explore ways to augment human decision-making processes with AI-driven insights to maximize the benefits of these technologies in medicine.

## Data Availability

All data produced in the present study are available upon reasonable request to the authors

